# A self-help fully automated digital intervention to improve subthreshold depressive symptoms among older adults in a socioeconomically deprived region of Brazil (PRODIGITAL): a pragmatic, two-arm randomised controlled trial

**DOI:** 10.1101/2023.11.20.23298686

**Authors:** Carina Akemi Nakamura, Nadine Seward, Tim J. Peters, Thiago Vinicius Nadaleto Didone, Felipe Azevedo Moretti, Marcelo Oliveira da Costa, Caio Hudson Queiroz de Souza, Gabriel Macias de Oliveira, Monica Souza dos Santos, Luara Aragoni Pereira, Mariana Mendes de Sá Martins, Pepijn van de Ven, William Hollingworth, Ricardo Araya, Marcia Scazufca

## Abstract

**Background:** Subthreshold depression is a risk factor for major depression and is associated with increased morbidity and mortality, especially in poorly resourced settings. There is emerging evidence that digital interventions may be effective at improving depressive symptoms in High Income Countries but not Low-and Middle-Income settings. We aimed to evaluate the effectiveness of the Viva Vida digital intervention at improving symptoms of subthreshold depression among older adults in socioeconomically deprived settings in Brazil.

**Methods:** PRODIGITAL was a single blind, two-arm, individually randomised controlled trial conducted in 46 primary care clinics in Guarulhos, Brazil. Individuals aged 60+ years were randomly contacted by phone for a screening assessment. Those who presented with anhedonia and/or depressed mood (Patient Health Questionnaire (PHQ)-2), and who subsequently scored between 5 and 9 on the PHQ-9 were invited to participate. The intervention (Viva Vida) arm received a self-help fully automated programme sent via WhatsApp with no support from healthcare professionals. Forty-eight audio and visual messages based on psychoeducation and behavioural activation approaches were delivered over six weeks. The control arm received a single message with information about depression. The primary outcome was the PHQ-9 at three months’ follow-up. All primary analyses were performed according to allocated arm with imputed data. The trial is registered with ReBEC, RBR-6c7ghfd.

**Findings:** Participants were recruited between 8 September 2021 and 19 August 2022. Of the 454 participants enrolled, 223 were randomised to the intervention arm and 231 to receive the single message. A total of 385 (84·8%) completed the three-month follow-up assessment; no difference in mean PHQ-9 scores between the arms (adjusted difference: -0·61; 95% CI: -1·76, 0·53; *p*=0·290) was observed.

**Interpretation:** These results demonstrate that a fully automated digital programme did not help to improve subthreshold depressive symptoms amongst older adults. More research is needed to understand how such interventions can be adapted to reduce depressive symptoms as well as prevent against major depression in low-resourced settings.

**Funding:** São Paulo Research Foundation and Joint Global Health Trials

**Research in context:** Evidence before this study

We searched PubMed for randomised controlled trials on Oct 20, 2023, using the search terms (((((((digital) OR (internet)) OR (web)) OR (mobile)) OR (e-health) OR (technology) AND (((subthreshold) OR (subclinical)) OR (subsyndromal))) AND (((depression) OR (depressive)))) AND (((older) OR (elderly) OR (senior))). We did not apply any restrictions on languages or date of publication. Two studies assessing a fully automated digital intervention for subthreshold depression were found. An internet-based cognitive behavioural therapy (iCBT) was delivered to adults aged 50+ years in the Netherlands, whilst automated emails using self-help behaviour strategies (Mood Memos) was sent to adults from 18 to 78 years (mean age of 36 years) in Australia. The interventions showed improvement on depressive symptoms at ten (iCBT) and six weeks (Mood Memos). Like the Viva Vida, one study conducted with adults in China used a messaging application (WeChat). However, WeChat offers more features, and the iCBT programme was guided by clinical psychologists.

**Added value of this study:** We report a self-help fully automated digital psychosocial intervention for subthreshold depression among older adults in Brazil. To the best of our knowledge, this is the first psychosocial intervention of this kind in a low-and middle-income country. The intervention was designed to be user-friendly and accessible to a population with low literacy and limited digital skills. While a significant portion of participants engaged with the programme by opening most of the messages, we did not observe any difference in improving depressive symptoms.

**Implications of all the available evidence:** Fully automated digital interventions with no support of health or lay professionals are usually low cost and can easily be scaled up. Consequently, they have the potential to improve access to care and help reduce the mental health treatment gap in an affordable way. When tailored to socioeconomically vulnerable individuals, these interventions can also help promote more equitable access to care. Future studies should focus on understanding how to adapt such interventions to be effective for subthreshold depression.

## Introduction

Depression is widely recognised as a leading cause of disability globally,^1^ and poses the largest burden of disease among all mental health conditions. Although depression is common across the life course, older adults are at increased risk of disability, increased mortality, and poorer outcomes from physical health.^2^ Depression has also been reported as a potential target for interventions due to its central role in multimorbidity in older populations.^3^

Subthreshold depression is defined as having one core symptom of depression, such as anhedonia and depressed mood, along with mild depressive symptoms that do not meet the criteria for major depressive disorder.^4,5^ Research suggests that subthreshold depression is an important risk factor for major depression. Indeed, studies indicate that between 10% and 50% of people with subthreshold depression develop major depression.^4^ As with major depression, older adults with subthreshold depression are at greater risk of worsening symptoms of comorbidities, functional decline, increased risk of mortality, and increased healthcare costs.^6^ Thus, there is an urgent need to develop and evaluate scalable interventions to manage subthreshold depression and prevent major depressive disorder developing in older adults, especially in low-and middle-income countries (LMICs) where the need is greatest.

The use of digital interventions to manage depressive symptoms has become increasingly popular.^7^ There is growing evidence that digital interventions may help to improve symptoms of subthreshold and major depression.^8^ However, studies in poorly resourced settings in LMICs are limited. Importantly, there is limited evidence on the effectiveness of these interventions in older individuals, increasing healthcare disparities.^9^ Given these interventions can be both low cost and easily scalable, they are potentially an important strategy to reduce the mental health gap in LMICs.^10,11^

We designed and evaluated a self-help fully automated digital psychosocial intervention (Viva Vida) for the management of depression among older adults in deprived regions of Brazil (PRODIGITAL-D).^12^ Findings from this research suggest that Viva Vida improved recovery from depression (9-item Patient Health Questionnaire (PHQ-9) scores <10) at the three-month follow-up, but no such evidence exists for the Viva Vida programme specifically tailored for older adults with subthreshold depression. Here we evaluate the effectiveness of this programme in the management of subthreshold depression among older adults in Guarulhos, Brazil (PRODIGITAL).

## Methods

### Study design and participants

PRODIGITAL was a two-arm, single blinded, individually randomised controlled trial (RCT) with a 1:1 allocation ratio. The study took place in Guarulhos, a municipality in the metropolitan region of São Paulo city with 1·3 million inhabitants (of which 13·8% is 60 years or over). The Family Health Strategy (FHS) is the main primary care model in Brazil and covers 31% of the Guarulhos population, mainly living in the most socioeconomically vulnerable areas. While this model aims to provide a more comprehensive care through multiprofessional teams with preventive and health promotion approaches, the traditional primary care model relies mostly on medical appointments to deliver primary care assistance. In this study, we included participants registered with 46 primary care clinics, known as Unidade Básica de Saúde (UBS), of which 37 were based on the FHS model and the remaining were a mix of that and the traditional primary care model.

Inclusion criteria were older adults (60+ years), access to WhatsApp, and subthreshold depressive symptomatology. The latter criterion was assessed using the PHQ and defined as at least one core symptom of depression^4^ (PHQ-2 score≥1), and mild depressive symptoms (PHQ-9 score between 5 and 9).^13^ Participants were also required to be registered with one of the participating UBSs. In the UBSs based on the mixed models, only individuals receiving care from the FHS were eligible. Exclusion criteria included significant vision or hearing impairment (not being able to listen or see messages received in the mobile phone), significant cognitive impairment or language barriers (not being able to understand the instructions or questions during the assessment), participants of our previous RCT (PROACTIVE),^14^ and another household member already enrolled in the study. Participants who answered positively to item-9 of the PHQ-9 (suicidal ideation) and were deemed as high suicidal risk by a standardised questionnaire were also excluded.

Verbal consent was granted for the screening assessment and for participation in the trial. This study was authorised by the Secretaria da Saúde do Município de Guarulhos and approved by the Ethics Committee of the Hospital das Clínicas da Faculdade de Medicina da Universidade de Sao Paulo – HCFMUSP (CAPPesq, ref: 4.683.191).

### Randomisation and masking

Participants were stratified by age (60-69, 70-79 years, 80+ years), gender (male/female), and type of UBS (full FHS/mixed models). The randomisation sequence was generated using random permuted blocks with random block sizes of six, eight or ten by researchers not directly involved in the data collection (CAN and TJP). The ‘randomization module’ of the Research Electronic Data Capture (REDCap)^15^ was used to conceal the randomisation sequence and randomise participants in the RCT.

Independent research assistants responsible for collecting data were masked to trial allocation, and two different teams were assigned for either the first or the second follow-up assessment. Also, whenever possible, research assistants did not perform more than one interview (baseline and follow-up) with the same participant. It was not possible to mask researchers responsible for scheduling the automated delivery of messages but they had no role in collecting data. Trial participants were not masked due to the nature of the intervention.

### Procedures

Screening for eligibility was conducted simultaneously with the PRODIGITAL-D RCT that recruited older adults with depressive symptomatology (PHQ-9 score≥10).^12^ An alphabetical list of the names and phone numbers of all individuals aged 59+ years registered with the eligible UBSs and receiving care from the FHS were provided by Guarulhos Health Secretary. Duplicate entries, individuals with no phone numbers, and PROACTIVE RCT participants were excluded. The remaining individuals received a random ID number (due to the alphabetical ordering of potential participants) when entered into the REDCap system. Details of the recruitment process have been published elsewhere.^16^

A pre-screening phase checked if the mobile phone numbers were valid and registered with WhatsApp. Using the randomly ordered list of ID numbers, those with a valid WhatsApp number were contacted by phone for the screening assessment. Individuals were first screened for inclusion and exclusion criteria, including the PHQ-2.^17^ If anhedonia or depressed mood was present (PHQ-2 scores≥1), then the remaining seven questions of the PHQ-9^13^ were asked. Those who scored between 5 and 9 on the PHQ-9 were considered to have subthreshold depressive symptoms^18^ and completed a baseline questionnaire regarding other mental and physical health conditions, quality of life, and sociodemographic profile. The screening assessment and baseline questionnaire took approximately 35 minutes.

Individuals who completed the baseline assessment were invited to participate in the RCT. During the invitation to participate, research assistants provided all information about the trial that took approximately 15 minutes. If the participants were not able to spend this extra time, this was completed in an additional call no more than 28 days after the PHQ-9 assessment. All assessments were conducted by phone and the calls were recorded if authorised by the individual.

Following consent, participants were randomised to either intervention or control arms. Two lists (one for the intervention and one for the control arms) were sent at the end of each week to the team responsible for sending the Viva Vida messages. The first message (intervention arm) and the single message (control arm) were sent out no later than ten days after consenting to participate in the trial. Participants in both arms continued to receive the usual care provided by the UBSs, and the research team did not interfere in any clinical decisions (medications, consultations, and treatments). Although UBS staff were not involved in the trial, the first message sent to participants in the intervention arm and the single message sent to participants in the control arm encouraged people to seek healthcare if they felt this was needed.

Participants in the intervention arm received a version of the Viva Vida programme developed for older adults with subthreshold depression. The programmes for depression and subthreshold depression have the same structure, but they differ on the content of the messages (how the story is told), and the intensity of the programme (based on the number of audio messages). Viva Vida for subthreshold depression is a six-week fully automated digital psychosocial intervention based on psychoeducation and behavioural activation. The programme was specifically designed for older adults with low levels of education and consisted of 48 brief audio and visual messages delivered by WhatsApp four days a week (twice a day). Audio messages using storytelling techniques were delivered in the morning. The three minutes audio message introduced a topic that was reinforced by a visual message (image with a short text) sent in the afternoon. Topics covered during the programme were about depressive symptoms and ways to deal with them, including adding more pleasant and/or meaningful activities in their daily lives. An extra message at the beginning of the programme informed participants to contact the technical support in case they have issues receiving the messages. In addition, once a week participants received a message with a question asking their opinion and/or experiences with the programme. Participants were able to answer the question using the WhatsApp ‘quick reply’ tool, in which up to three pre-defined responses were offered. Participants were also invited to send a text or audio message to complement or justify their answers. All spontaneous messages sent by participants were replied with an automated message.

Participants in the control arm received one audio message that was approximately six minutes long. The message addressed depressive symptoms, provided guidance on their management, and included tips for maintaining a healthier lifestyle.

A dedicated web system integrated into the WhatsApp Business Application Programming Interface (API) was developed to schedule and deliver the messages. The system is hosted on a cloud service that is encrypted through Secure Socket Layer certificates. The API collected and managed WhatsApp data, including timestamps (date and time) for when messages were sent, delivered, and ‘read’ (that is, the chat window was opened). Timestamps and the answers were also provided when participants answered the weekly questions using the ‘quick reply’ tool.

During the first two weeks of the programme, the research team monitored the status of the messages to identify and contact participants with technical issues (such as those who had not received or opened the messages). This allowed us to identify and correct any potential issues with our system, or the WhatsApp Business app, ensuring that participants received the messages as intended. After this period, no additional contact was made from the research team until the follow-up assessments.

Follow-ups were completed by phone at three months (weeks 12 to 16) and five months (weeks 20 to 24) after receiving the first message. A four-week window was used to maximise response to follow-up considering the reality of conducting phone interviews. Information collected in the assessments included measures on depressive symptomatology (PHQ-9), anxiety symptomatology assessed using the Generalised Anxiety Disorder-7 (GAD-7),^19^ perceived loneliness assessed using the 3-item University of California, Los Angeles (UCLA) loneliness scale (3-item UCLA),^20^ health-related quality of life with the European Quality of Life five-dimensional questionnaire, five-level version (EQ-5D-5L),^21^ and capability wellbeing assessed with the ICEpop CAPability measure for Older people (ICECAP-O).^22^ All assessment data were collected and managed using REDCap^15^ hosted at the Hospital das Clínicas da Faculdade de Medicina da Universidade de Sao Paulo. Quality control of a random sample of recorded assessments was conducted by an independent research assistant (not involved in any data collection) to ensure all procedures have been followed and the quality of data collected.

Severe adverse events, including acute suicidal risk, hospitalisations and death were collected during the follow-up assessments for participants in both study arms. Participants who reported suicidal ideation were assessed for acute suicidal risk by a standardised protocol used in a previous study.^14^ UBS managers were contacted to inform about participants with suicidal ideation. Hospital admissions and death were investigated with participants and/or family members to understand if it was related to study participation.

### Outcomes

The primary outcome was self-reported depressive symptoms captured using the PHQ-9 score at three months. This outcome was also assessed at five months as a secondary outcome. Additional secondary outcomes measured at both three and five months included: depressive symptomatology defined as PHQ-9 score≥10,^13^ anxiety symptomatology (GAD-7),^19^ loneliness (3-item UCLA),^20^ health-related quality of life (EQ-5D-5L),^21^ and capability wellbeing (ICECAP-O).^22^

### Sample size

We aimed for a large and clinically significant difference in depressive symptoms between the intervention and control arms of 0·33 standard deviations in PHQ-9 scores.^23^ A sample size of 142 to 162 individuals in each arm (284 to 324 in total) can detect this difference at three months with 80% to 85% power with a two-sided 5% significance level. Albeit for a different study group and intervention, our PROACTIVE pilot study indicated that this effect size is feasible with psychosocial interventions (PHQ-9 means of 5.9 and 6.4 with a standard deviation of 1.5; data not published). We anticipated 25% attrition and therefore aimed to recruit 225 individuals in each arm for a total sample size of 450.

### Statistical analysis

#### Primary and secondary outcomes

The primary and secondary outcomes of difference in mean PHQ-9 scores between the intervention and control arms at three month and five months respectively, were evaluated using linear regression models that assume residuals are normally distributed. The secondary outcomes of difference in mean GAD-7 scores, EQ-5D-5L scores, ICECAP-O scores, and 3-item UCLA scores captured at three and five months, were also evaluated using linear regression models. The secondary outcome of risk of depressive symptomatology (defined as PHQ-9≥10) at three and five months was evaluated using fixed effects Poisson regression models with a log-link function and robust error variances. All regression analyses were adjusted for stratification (age group, gender, and type of UBS) and baseline PHQ-9 scores (plus baseline values of the corresponding outcome for analyses of all outcomes not based on PHQ-9 scores).

For each arm separately, missing data for all outcomes were replaced using multiple imputation by chained equations (MICE) as implemented in the MI command in Stata 17 (StataCorp) under the assumption that data were missing at random (MAR). Supplementary material provides details of the methods used for the missing data analyses including sensitivity analyses as well as results comparing estimates using complete case versus imputed data.

### Subgroup analyses

Pre-specified subgroup analyses were investigated using the Wald test for interactions assessed at both follow-up assessments using product terms between the treatment arm and the following variables: gender; age; educational level; comorbid physical illness (diabetes, hypertension, and both); and baseline PHQ-9 levels.

### CACE analyses

Complier Average Causal Effect (CACE) analysis^24^ using an instrumental variable estimator and imputed data was applied to estimate the effect of the number of messages electronically recorded as being ‘opened’ on depression outcomes. The protocol^12^ and the SAP pre-specified the ‘minimum therapeutic dose’ as listening to “most of the messages”, as it was initially based on self-reported categories (“none”, “a few”, “at least half”, “most of the messages”, and “all of the messages”) of a question from the first follow-up assessment. In the event, we were able to access the electronic data, and we therefore operationalized the pre-specified minimum therapeutic dose as ‘opened’ 36 or more messages (75% of the total of 48 messages) versus ‘opening’ 35 or fewer messages. The CACE analyses were conducted using PHQ-9 score at three and five months, adjusting for stratification. We also conducted sensitivity analyses using thresholds of ‘opening’ at least half of the total messages (24+) versus 23 messages or fewer, and ‘opening’ all 48 messages versus not doing so.

### Sensitivity analysis

Any baseline variables imbalanced substantially between treatment arms (assessed using descriptive statistics) were adjusted in a secondary analysis for the primary outcome. We also performed a sensitivity analysis for the primary outcome adjusting for the time between randomisation and the primary follow-up assessment at three months. For both these sensitivity analyses, if there was a noticeable impact, we planned to conduct similar analyses for the secondary outcomes.

Regression diagnostics were run for both the linear and Poisson regression models. Normality assumptions for all linear regression models were evaluated by examining the residual plots. Stata’s post estimation command *gof*, was used to assess the Poisson model.

Statistical tests were two-sided, and all analyses were conducted using Stata 17 (StataCorp). The trial was registered with the ReBEC, RBR-6c7ghfd.

## Results

Recruitment of participants started on 8 September 2021 and ended on 9 August 2022, with follow-up assessments completed on 30 January 2023. A total of 50,351 individuals registered with 46 UBSs entered our system, of which 22,484 (44·7%) had a valid phone and WhatsApp access and were randomly screened for eligibility by a phone call. Among those who were excluded after at least two contact attempts, 5,306 (24·1%) were contacted and did not meet the inclusion criteria, 1,144 (5·2%) declined to participate, two (0·01%) lived in the same household as another participant of the RCT, and 15,578 (70·7%) were not found. Hence, 454 participants were recruited, 223 (49·1%) randomised to the intervention arm and 231 (50·9%) to the control arm (Figure 1).

**Figure 1:**
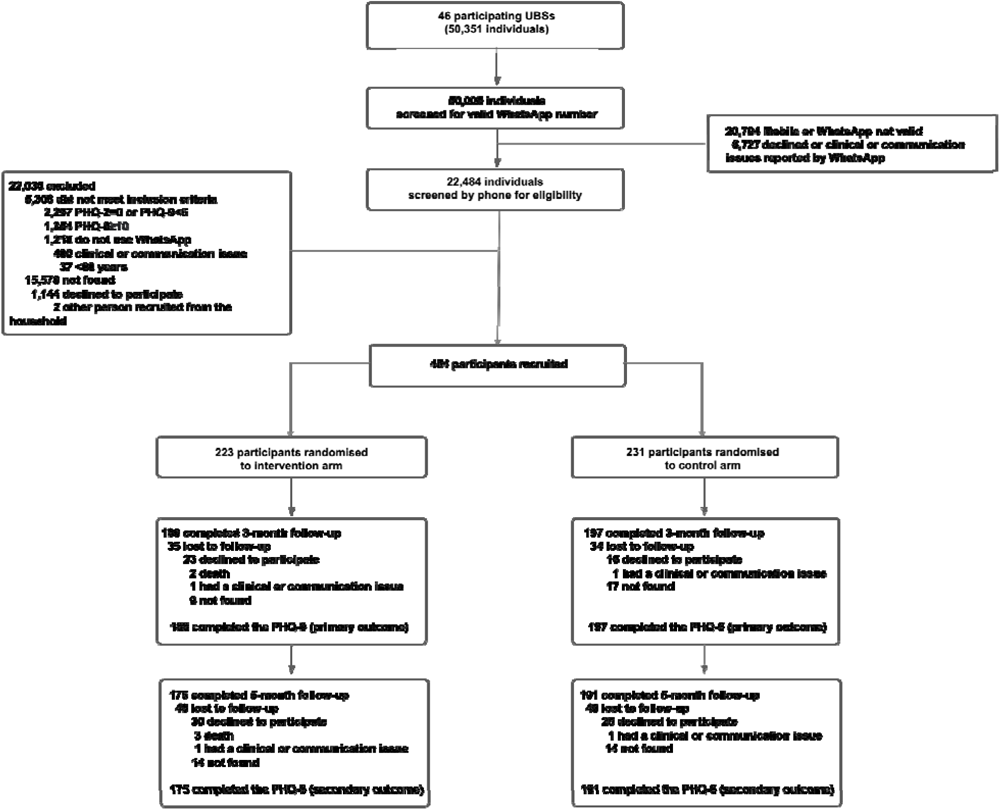
Trial profile.

A total of 385 (84·8%) participants recruited were followed up at the three-month assessment. Similar proportions of participants were lost to follow-up in the intervention (n=35, 15·7%) and control arms (n=34, 14·7%). At the five-month follow-up assessment, 366 (80·6%) participants were followed up, with a greater proportion lost to follow-up at five months in the intervention arm (n=48, 21·5%) compared with the control (n=40, 17·3%) arm.

Descriptive statistics of baseline characteristics indicate most of the participants were 60–69- year-old women, of whom the majority had less than eight years of education and were on the minimum wage (Table 1). This table indicates some potential imbalances between arms, with for instance slightly more participants in the control arm reporting hypertension and diabetes, and slightly fewer reporting taking antidepressants.

Sixteen participants (7·2%) in the intervention arm and ten (4·3%) in the control arm reported hospitalisation not related to mental health during the trial. Three (1·3%) deaths were reported, all in the intervention arm. None of the events were considered related to participation in the trial.

### Primary and secondary outcomes

The results from the primary analysis demonstrated no substantial difference in mean PHQ-9 scores at three months in the intervention arm (6·8 [SD: 5·5]) compared with the control arm (7·1 [SD: 5·6]) (Table 2). The adjusted difference in mean PHQ-9 scores after imputing missing values was -0·61 (95% confidence interval (CI): -1·76, 0·53; *p*=0·290), and in terms of units of standard deviation all values in the CI are below our target threshold of 0.33. At the five-month follow-up and after imputing missing data, the adjusted difference in mean PHQ-9 scores in the intervention versus the control arm also demonstrated a lack of effect of the intervention (-0·47 [95% CI: -1·6, 0·7]; *p*=0·425). These findings were similar to those from the complete case analysis (Supplementary Material).

There was no evidence of effects of the intervention on any of the secondary outcomes including proportion of participants with depressive symptomatology (PHQ-9≥10), anxiety symptomatology (GAD-7 scores), loneliness (3-item UCLA score), EQ-5D-5L quality of life, and ICECAP-O capability scores in both the complete case analyses (Supplementary Material) and using imputed data (Tables 3 and 4).

### Subgroup analyses

Findings from our subgroup analyses (Supplementary material) revealed that there was evidence of an interaction between gender and treatment arm (*p*<0·028). Using the post estimation *lincom* command in Stata, there was a notable difference between mean PHQ-9 scores at the three-month follow-up in favour of the intervention for women (adjusted difference in means: -1·46 [95% CI: -2·85, -0·08], with no such effect amongst the men (xx [yy, zz]).

### CACE analyses

The electronic system indicated that 177 (79·4%) participants in the intervention arm ‘opened’ at least 36 of the 48 messages (75% of the total messages; our pre-specified minimum therapeutic dose). Estimates from the upper and lower thresholds of our sensitivity analyses indicates that 186 (83·4%) participants ‘opened’ at least 24 messages (50% of the total messages), and 132 (59·2%) participants ‘opened’ all 48 messages. Estimates from the CACE analysis (Table 5) evaluating the effect of opening at least 36 messages, compared with participants opening less than this amount (the pre-specified ‘minimum therapeutic dose’) after imputing missing data did not show an improvement in mean PHQ-9 scores, at both three-month (adjusted difference between means: -0·93 [95% CI: -2·32, 0·46]; *p*=0·191) and five-month assessments (-0·75 [95% CI: -2·21, 0·71]; *p*=0·313). The sensitivity analyses testing upper and lower limits of opening messages came to the same conclusions.

### Sensitivity analyses

Given the imbalances between the treatment arms, we ran a sensitivity analysis adjusting liberally for them as well as stratification factors that were accounted for in the analyses of our primary outcome. This sensitivity analysis for our primary outcome suggests that estimates shifted towards the null, but the 95% CIs widened and still overlapped substantially with those from the primary analysis (Supplementary Material). Adjusting for the number of days between randomisation and the first follow-up visit led to no appreciable impact on the results for the primary outcome (Supplementary material), and hence neither of these sensitivity analyses were extended to the secondary outcomes.

## Discussion

To the best of our knowledge Viva Vida is the first self-help fully automated digital psychosocial intervention aiming to manage subthreshold depression among older adults in LMICs. Viva Vida delivered automated messages based on psychoeducation and behavioural activation without any support from trained healthcare workers. Our findings suggest that Viva Vida did not demonstrate any improvement in depressive symptoms among older adults at three and five months. Nor did Viva Vida prevent participants developing major depression at either follow-up assessments. Results also suggest there were no improvements in any of our other secondary outcomes (anxiety symptomatology, perceived loneliness, health-related quality of life, and capability wellbeing). Interestingly, our subgroup analyses indicated that compared with men, women experienced a greater reduction in symptoms of depression at the three-month assessment.

Previous studies showed evidence of the effectiveness of digital interventions for subthreshold depression among adults and older adults.^25^ However, many of these studies offered some kind of support or contact with a health professional and the majority were conducted in high-income countries. We identified only two fully automated digital interventions without professional support for individuals with subthreshold depression, but both included younger people, and the content and techniques differed from PRODIGITAL. One study assessing an eight-week intervention based on cognitive behavioural therapy found a moderate effect size in participants over 50 years old (mean age of 55 years) in the Netherlands.^26^ Another study delivered automated emails with self-help behaviour strategies (Mood Memos) over six weeks to adults (mean age of 36 years) in Australia.^27^ This intervention showed a small effect size in reducing PHQ-9 scores but no differences on the risk of major depression.

One potential explanation for our null findings is the absence of contact with professionals or peers, which has been reported as an important feature by authors of a systematic review of digital interventions for older adults.^7^ Our programme was fully automated, whereby participants only interacted with a ‘quick reply’ tool. Standardised and automated messages were then sent to participants based on their responses. However, lack of human interaction may not be the only reason that helps to explain our null findings. Indeed, the Viva Vida programme that we designed for participants with depression (PHQ-9 score≥10) was effective at three months. Given participants in the subthreshold depression study received a lower proportion of psychoeducation or behavioural activation audio messages (80% versus 50%) it is possible that a slightly more intense programme may be better suited for improving subthreshold depression. Future studies could consider adapting the Viva Vida programme to include cost effective solutions to provide social support as well as increasing the number of sessions offered to participants.

This Viva Vida digital programme was especially tailored to the target population, considering low levels of education and socioeconomic condition, to ensure the acceptability, appropriateness, and feasibility to older adults. To accommodate those with lower literacy levels, we incorporated audio and visual messages with minimal written content. Moreover, the programme’s characters used an appropriate and simple language to share stories that older adults could identify with, encouraging reflection on similar life situations. Simpler and user-friendly technologies were identified by the previously described systematic review as another important feature of successful digital intervention for older adults.^7^ WhatsApp is the most used messaging application in Brazil, and it is popular including among this age group.^28^ This design choice eliminated the need for participants to download new applications or acquire additional digital skills. We believe these elements contributed to a positive overall experience reported by participants; findings that will be described in a publication of our process evaluation.

The lack of a well-established standardised criteria to capture subthreshold depression is an important consideration for future research. While this study assessed subthreshold depressive symptoms to recruit participants using a validated screening scale that could be easily used in primary care, other studies that assessed psychosocial interventions for older adults with subthreshold depression used standardised clinical assessments.^23,29^ Additionally, unlike these studies that utilised the PHQ-9 score or other validated screening scales solely to assess the outcome measure, we employed the same instrument for both eligibility criteria and the primary outcome assessment. Given initial symptoms severity is known to influence reduction in symptoms of depression,^30^ it is difficult to compare findings with other studies. Consideration is needed to standardise definitions of subthreshold depression, so results from different trials are comparable.

Subgroup analyses suggested that women were more likely to benefit from the Viva Vida programme than men. Besides most participants were women. Gender differences should also be considered when designing a digital intervention. Future studies should explore the potential benefits of personalised approaches based on gender, as well as other relevant socioeconomic or clinical characteristics (for example, education level or comorbidities), and individual preferences to enhance the programme’s effectiveness and overall experience.

This study is not without limitations. An important consideration is our system’s ability to accurately monitor adherence to the intervention. Information provided by WhatsApp was the ‘read’ receipts, indicating if/when the chat window was open and only if the feature was not turned off by the participant. Therefore, it was not possible to evaluate the actual adherence (listening to the messages) to the intervention. Importantly, recruitment was conducted over the phone and one inclusion criterion was ensuring participants had access to WhatsApp. Although this is a popular application among older adults in Brazil, it is possible that the trial may have unintentionally created a digital divide whereby the most socioeconomically vulnerable individuals were excluded. This could have influenced our findings by over or underestimating the treatment effect. As an example, vulnerable participants may be more or less likely to respond to the digital intervention.

Self-help fully automated digital interventions offer a low-cost and highly scalable approach, that is particularly valuable in LMICs with limited mental healthcare resources. Tailored digital interventions for older adults are essential to provide a better experience and ensure access to care for this population. Future research should focus on adapting and improving this programme, given its demonstrated effectiveness for more severe depressive symptoms. Consideration should also be given as to whether fully automated digital programmes can be used as part of a stepped-care strategy to treat and prevent the development of depression in older adults.

## Data Availability

Data will be available for reasonable requests from authors approximately one year after publication

