## Supplementary tables for "A self-help fully automated digital intervention to improve subthreshold depressive symptoms among older adults in a socioeconomically deprived region of Brazil (PRODIGITAL): a pragmatic, two-arm randomised controlled trial"

Sensitivity analyses - missing data analyses

Methods

Initially, patterns of missingness were investigated by comparing missing with complete data for the baseline demographic characteristics and morbidity questionnaires with mean PHQ-9 scores, by trial arm, at both follow-up visits. In order to reduce bias and loss of information, we used multiple imputation by chained equations (MICE) with 50 imputations, as implemented in the MI command in Stata under the assumption that data were missing at random (MAR).^1^ Variables used in the MICE models consisted of the outcome of PHQ-9 scores, covariates described in the Statistical Analysis Plan (age group, baseline PHQ-9 score, gender, treatment arm, and type of primary care centre) and variables found to be predictors of missingness.^2, 3^ Predictors of missingness include the following: age group at baseline, health-related quality of life (EQ-5D-5L) at baseline, and tobacco use at baseline. Potential predictors that theoretically could have influenced levels of missingness include personal income at baseline, and levels of enjoyable activities participants engaged in. Baseline scores for corresponding outcome models including depressive symptomatology (PHQ-9), anxiety symptomatology (GAD-7), capability wellbeing (ICECAP-O), and levels of loneliness captured with 3-item UCLA scores were also included.

To assess the sensitivity of our findings against modest departures from the MAR assumption, a weighted sensitivity analysis using the Selection Model Approach was applied.^4-6^ Briefly, once data had been imputed under MAR, parameter estimates from each imputed dataset were reweighted to allow for the data to be missing not at random (MNAR). The chosen weights, used to reweight the data to account for MNAR, are dependent on the assumed degree of departure from MAR. The parameter used to re-weight the data, denoted by δ, is the log odds ratio of the probability of not being lost to follow-up and data on depression outcomes present, when participants were depressed, compared to when a participants were not depressed.^4-6^ If δ=0, losses to follow-up and therefore the outcome of risk of depression being not missing, could be considered to be MAR. δ>0 indicates that the probability of not being lost to follow-up and the depression outcome present, when depression was present, was greater than when depression was absent. δ<0 indicates that the probability of not being lost of follow-up when depression was present was less. As δ decreases from zero, the probability of participants not being lost to follow-up when depression was present is less than the probability of participants not being lost to follow-up when participants were not depressed (i.e. greater probability of being lost to follow-up depression was not present).

We hypothesize it is more likely that participants being lost to follow-up and our outcome data being missing in instances where depression was not present, compared to when depression was present (i.e. δ>0). To test the stability of our model, we considered different degrees of departure from the MAR assumption by considering plausible values of δ ranging from 0.10 to 0.40. This range corresponds to risk of depression for the data being observed when a participant had depression compared to when it did not, ranging from 0.10 to 0.40.

Results

General

eTables 1 and 2 demonstrate findings from the analysis comparing baseline demographics and baseline measures for the secondary outcomes between participants with complete data, and those with missing data, at three- and five-months follow-up visits respectively.

eTable1: Comparison between participants with complete data and participants with missing data at the three-month follow-up visit

| **Number/total sample size (%)** | | | | |
| --- | --- | --- | --- | --- |
|  | **Control** | | **Viva Vida** | |
|  | **Total (n=197)** | **Missing PHQ-9 (n=34)** | **Total (n=188)** | **Missing PHQ-9 (n=35)** |
| **Tobacco**  No | 204 (88.3)  27 (11.7) | 31 (91.2)  3 (8.8) | 177 (79.4)  45 (20.6) | 21 (60.0)  14 (40.0) |
| Yes |  |  |  |  |
| **Gender, No. (%)** |  |  |  |  |
| Male | 82 (35.5) | 10 (29.4) | 80 (35.9) | 14 (40.0) |
| Female | 149 (64.5) | 24 (70.6) | 143 (64.1) | 21 (60.0) |
| **Age group, No. (%)** |  |  |  |  |
| 60-69 years | 183 (79.2) | 25 (73.5) | 181 (81.2) | 33 (94.3) |
| 70+ years | 48 (20.8) | 9 (26.5) | 42 (18.8) | 2 (5.7) |
| **Education, No. (%)** |  |  |  |  |
| No | 29 (12.6) | 2 (5.9) | 20 (8.9) | 4 (11.4) |
| Yes | 202 (87.5) | 32 (94.1) | 203 (91.0) | 31 (88.6) |
| **Education (level), No. (%)** |  |  |  |  |
| None | 35 (15.2) | 3 (8.8) | 26 (11.7) | 5 (14.3) |
| 1-4 years | 64 (27.8) | 10 (29.4) | 48 (21.5) | 10 (28.6) |
| 5-8 years | 47 (20.4) | 11 (32.4) | 53 (23.8) | 7 (20.0) |
| >8 years | 84 (36.5) | 10 (29.4) | 96 (43.1) | 13 (37.1) |
| **Job, No. (%)** |  |  |  |  |
| No | 149 (64.5) | 20 (58.8) | 140 (62.8) | 21 (60.0) |
| Yes | 82 (35.5) | 14 (41.2) | 83 (37.2) | 14 (40.0) |
| **Personal income, No. (%)** |  |  |  |  |
| Up to 1 MW | 118 (51.3) | 18 (52.9) | 123 (51.2) | 16 (48.5) |
| >1-2 MW | 56 (24.4) | 8 (23.5) | 50 (22.8) | 10 (30.3) |
| >2 MW | 56 (24.4) | 8 (23.5) | 7 (21.2) | 7 (21.2) |
| **Household income, No. (%)** |  |  |  |  |
| Up to 1 MW | 31 (17.7) | 3 (14.3) | 28 (17.8) | 5 (18.5) |
| >1-2 MW | 57 (32.6) | 9 (42.9) | 44 (28.0) | 9 (33.3) |
| >2 MW | 87 (49.7) | 9 (42.9) | 85 (54.1) | 13 (48.2) |
| **Hypertension (self-reported), No. (%)** |  |  |  |  |
| No hypertension | 75 (32.5) | 12 (35.3) | 84 (37.7) | 10 (28.6) |
| Hypertension | 156 (67.5) | 22 (64.7) | 139 (62.3) | 25 (71.4) |
| **Diabetes (self-reported), No. (%)** |  |  |  |  |
| No diabetes | 148 (64.1) | 23 (67.7) | 155 (69.5) | 25 (71.4) |
| Diabetes | 83 (35.9) | 11 (32.4) | 68 (30.5) | 10 (28.6) |
| **Mean PHQ-9 score (SD)** |  |  |  |  |
|  | 6.9 (1.4) | 6.8 (1.5) | 7.2 (1.4) | 6.9 (1.4) |
| **Number of days participant engaged in enjoyable, meaningful activities over the past two weeks, No. (%)** |  |  |  |  |
| Not at all | 33 (11.8) | 4 (11.8) | 37 (16.6) | 3 (8.6) |
| Several days | 38 (16.5) | 5 (14.7) | 40 (17.9) | 8 (22.9) |
| Over half the days | 43 (18.7) | 5 (14.7) | 37 (16.6) | 6 (17.1) |
| Nearly every day | 116 (50.4) | 20 (58.8) | 109 (48.9) | 18 (51.4) |

*Abbreviations: MW: minimum wage (in 2021, the minimum wage in Brazil was BRL1110 (approximately US$213); PHQ-9: 9-item Patient Health Questionnaire for depression.*

eTable 2: Comparison between participants with complete data and participants with missing data at the five-month follow-up visit

| **Number/total sample size (%)** | | | | |
| --- | --- | --- | --- | --- |
|  | **Control arm** | | **Viva Vida** | |
|  | **Total (n=191)** | **Missing PHQ-9 (n=40)** | **Total (n=175)** | **Missing PHQ-9 (n=48)** |
| **Smoker, No. (%)** |  |  |  |  |
| No | 204 (88.3) | 36 (90.0) | 177 (79.4) | 31 (64.6) |
| Yes | 27 (11.7) | 4 (10.0) | 46 (20.6) | 17 (35.4) |
| **Gender, No. (%)** |  |  |  |  |
| Male | 82 (35.5) | 14 (35.5) | 80 (35.9) | 17 (35.4) |
| Female | 149 (64.5) | 26 (65.0) | 143 (64.1) | 31 (64.6) |
| **Age group, No. (%)** |  |  |  |  |
| 60-69 years | 183 (79.2) | 28 (70.0) | 181 (81.2) | 41 (85.4) |
| 70+ years | 48 (20.8) | 12 (30.0) | 42 (18.8) | 7 (14.6) |
| **Education, No. (%)** |  |  |  |  |
| No | 29 (12.6) | 2 (5.0) | 20 (9.0) | 6 (12.5) |
| Yes | 202 (87.5) | 38 (95.0) | 203 (91.0) | 42 (87.5) |
| **Education (level), No. (%)** |  |  |  |  |
| None | 35 (15.2) | 3 (7.5) | 26 (11.7) | 7 (14.6) |
| 1-4 years | 64 (27.8) | 64 (27.8) | 48 (21.5) | 13 (27.1) |
| 5-8 years | 47 (20.4) | 47 (20.4) | 53 (23.8) | 11 (22.9) |
| >8 years | 84 (36.5) | 84 (36.5) | 96 (43.1) | 17 (35.4) |
| **Job, No. (%)** |  |  |  |  |
| No | 149 (64.5) | 24 (60.0) | 140 (62.8) | 31 (64.6) |
| Yes | 82 (35.5) | 16 (40.0) | 83 (37.2) | 17 (35.4) |
| **Personal income, No. (%)** |  |  |  |  |
| Up to 1 MW | 118 (51.3) | 15 (37.5) | 123 (56.2) | 27 (58.7) |
| >1-2 MW | 56 (24.4) | 12 (30.0) | 50 (22.8) | 13 (28.3) |
| >2 MW | 56 (24.4) | 13 (32.5) | 46 (21.0) | 6 (13.0) |
| **Household income, No. (%)** |  |  |  |  |
| Up to 1 MW | 31 (17.7) | 3 (12.0) | 28 (17.8) | 6 (17.7) |
| >1-2 MW | 57 (32.6) | 9 (36.0) | 44 (28.0) | 14 (41.2) |
| >2 MW | 87 (49.7) | 13 (52.0) | 85 (54.1) | 14 (41.2) |
| **Hypertension (self-reported), No. (%)** |  |  |  |  |
| No hypertension | 75 (32.5) | 14 (35.0) | 84 (37.7) | 16 (33.3) |
| Hypertension | 156 (67.5) | 26 (65.0) | 139 (62.3) | 32 (66.7) |
| **Diabetes (self-reported), No. (%)** |  |  |  |  |
| No diabetes | 148 (64.1) | 25 (62.5) | 155 (69.5) | 33 (68.8) |
| Diabetes | 83 (35.9) | 15 (37.5) | 68 (30.5) | 15 (31.3) |
| **Mean PHQ-9 score (SD)** |  |  |  |  |
|  | 6.9 (1.3) | 6.9 (1.4) | 7.2 (1.4) | 7.1 (1.4) |
| **Number of days participant engaged in enjoyable, meaningful activities over the past two weeks, No. (%)** |  |  |  |  |
| Not at all | 33 (14.4) | 7 (17.5) | 37 (16.6) | 3 (6.3) |
| Several days | 38 (16.5) | 5 (12.5) | 40 (17.9) | 11 (22.9) |
| Over half the days | 43 (18.7) | 6 (15.0) | 37 (16.6) | 9 (18.8) |
| Nearly every day | 116 (50.4) | 22 (55.0) | 109 (48.9) | 25 (52.1) |

*Abbreviations: MW: minimum wage (in 2021, the minimum wage in Brazil was BRL1110 (approximately US$213); PHQ-9: 9-item Patient Health Questionnaire for depression.*

Sensitivity analysis checking the missing at random (MAR) assumption for the primary outcome

Results from the sensitivity analysis testing the MAR assumption for the secondary outcome of relative risk of depression at three months suggest that estimates moved towards the null (Participants with depression less likely to have missing data than participants without). At five months, findings suggest a similar mechanism for missing data.

eTable 3: Model 1 - Adjusted risk ration for the odds of having depression (95% CI) for different departures from the missing at random assumption, assuming greater probability of the outcome being missing when depression was present

| **Delta** | **Adjusted relative risk of depression (95% CI)** |
| --- | --- |
| 0.1 | 0.902 (0.585, 1.390) |
| 0.2 | 0.900 (0.585, 1.386) |
| 0.3 | 0.900 (0.586, 1.381) |
| 0.4 | 0.899 (0.587, 1.378) |

eTable 3: Model 2 - Adjusted risk ration for the odds of having depression (95% CI) for different departures from the missing at random assumption, assuming greater probability of the outcome being missing when depression was present

| **Delta** | **Adjusted relative risk of depression (95% CI)** |
| --- | --- |
| 0.1 | 0.667 (0.394, 1.129) |
| 0.2 | 0.652 (0.386, 1.101) |
| 0.3 | 0.636 (0.378, 1.070) |
| 0.4 | 0.621 (0.372, 1.039) |

**Results comparing estimates from MICE models with complete case analyses for both primary and secondary outcomes**

eTable 5: Comparison of estimates from multiple imputed data using MICE models, with estimates from complete case analysis for the primary outcome of mean difference in PHQ-9 scores between treatment arms and mean difference of other continuous secondary outcomes at three months

|  | **Complete case analysis^a,b^** | | **Imputed data from MICE analyses^a,b^** | |
| --- | --- | --- | --- | --- |
| **Model** | **Difference in means (95% CI)** | ***P* Value** | **Difference in means (95% CI)** | ***P* Value** |
| PHQ-9 scores^a^ | -0.64 (-1.75, 0.46) | 0.251 | -0.52 (-1.70, 0.65) | 0.380 |
| GAD-7 scores^a^ | -0.64 (-1.80, 0.52) | 0.277 | 0.57 (-1.64, 0.51) | 0.300 |
| EQ-5D-5L scores^a^ | -0.004 (-0.023, 0.015) | 0.705 | -0.004 (-0.024, 0.016) | 0.690 |
| ICECAP-O scores^a^ | 0.009 (-0.019, 0.036) | 0.545 | 0.011 (-0.017, 0.039) | 0.443 |
| 3-item UCLA scores^a^ | -0.25 (-0.55, 0.05) | 0.106 | -0.27 (-0.58, 0.04) | 0.084 |

^a^ Differences in means were estimated using linear regression models, adjusted for relevant baseline assessment of corresponding outcome, gender, age-group, type of primary care centre (Family Health Strategy and traditional primary care)

^b^ All estimates had missing data imputed seperately, by trial arm, using MICE models that included predictors of missingness, gender, age-group, type of primary care centre , and imbalances in missingness between treatment arms, and baseline value of

corresponding outcome variable.

PHQ-9 scores range from 0 to 27, with higher scores representing more severe symptoms of depression

GAD-7 scores range from 0 to 21 with higher scores representing more severe symptoms of anxiety

EQ-5D-5L scores range from -0.264 to 1, with higher scores representing higher quality of life

ICECAP-O scores range from 0 to 1, with higher scores representing greater levels of capability wellbeing

3-item UCLA scores range from 3 to 9, with higher scores representing more severe levels of loneliness

eTable 6: Comparison of estimates from multiple imputed data using MICE models, with estimates from complete case analysis for secondary outcomes at five months

|  | **Complete case analysis^a,b^** | | **Imputed data from MICE analyses^a,b^** | |
| --- | --- | --- | --- | --- |
| **Model** | **Difference in means (95% CI)** | ***P* Value** | **Difference in means (95% CI)** | ***P* Value** |
| PHQ-9 scores^a^ | -0.52 (-1.70, 0.65) | 0.380 | -0.43 (-1.59, 0.73) | 0.468 |
| GAD-7 scores^a^ | 0.15 (-1.06, 1.35) | 0.812 | 0.06 (-1.06, 1.19) | 0.912 |
| EQ-5D-5L scores^a^ | 0.007 (-0.011, 0.025) | 0.458 | 0.008 (-0.011, 0.026) | 0.414 |
| ICECAP-O scores^a^ | 0.009 (-0.020, 0.038) | 0.540 | 0.013 (-0.017, 0.042) | 0.407 |
| 3-item UCLA scores^a^ | 0.00 (-0.28, 0.28) | 0.998 | 0.02 (-0.31, 0.34) | 0.923 |

^a^ Differences in means were estimated using linear regression models, adjusted for relevant baseline assessment of corresponding outcome, gender, age-group, type or primary care centre (Family Health Strategy and traditional primary care)

^b^ All estimates had missing data imputed seperately, by trial arm, using MICE models that included predictors of missingness, gender, age-group, type of primary carer centre, and baseline value of corresponding outcome measure. Differences in missingness were also included in MICE models.

PHQ-9 scores, corresponding baseline value for outcome in question

PHQ-9 scores range from 0 to 27, with higher scores representing more severe symptoms of depression

GAD-7 scores ranges from 0 to 21 with higher scores representing more severe symptoms of anxiety

EQ-5D-5L scores range from -0.264 to 1, with higher scores representing higher quality of life

ICECAP-O scores range from 0 to 1, with higher scores representing greater levels of capability and general wellbeing

3-item UCLA scores range from 3 to 9, with higher scores representing more severe levels of loneliness

eTable 7: Comparison of adjusted relative risks from multiple imputed data using MICE models, with estimates from complete case analysis for the secondary outcomes of depression at three months and five months

|  | **Complete case analysis^a,b,c^** | | **Imputed data from MICE analyses^a,b,c^** | |
| --- | --- | --- | --- | --- |
| **Model** | **Odds ratio (95% CI)** | ***P* Value** | **Odds ratio (95% CI)** | ***P* Value** |
| Depression at three months^a,b^ (n=) | 0.93 (0.70, 1.24) | 0.634 | 0.96 (0.72, 1.27) | 0.762 |
| Depression at five months^a,b^ (n=) | 0.74 (0.50, 1,09) | 0.122 | 0.73 (0.50, 1.06) | 0.104 |

^a^ The primary and secondary outcome of recovery from depression was defined as PHQ-9 scores less than 10

^b^ Odds ratios and 95% CIs were calculated using Poisson regression models with a loglink function

c Models were adjusted for adjusted for gender, age-group, baseline PHQ-9 score and type of primary care centre (Family Health Strategy and traditional primary care).

^c^ All estimates had missing data imputed by trial arm using MICE models that included gender, age-group, type of primary care centre, baseline PHQ-9 score and predictors of missingness

Sensitivity analyses checking for maintenance of randomisation and subgroup analyses testing for moderation (complete cases only)

Sensitivity analysis to determine if randomisation was maintained at the three- and five-month follow-up visits

The comparison at the three-month visits (eTable 11) suggests a slight imbalance whereby participants in the intervention arm had higher levels of hypertension and a greater proportion of participants with more severe forms of depression, compared with participants in the control arm. Additionally, at the three-month follow-up visit participants in the control arm received a greater proportion of participants reporting pharmacological treatment for depression and a greater proportion of participants on higher incomes, than participants in the intervention arm. At the five-month follow-up visit (eTable 12), participants in the control arm had higher levels of hypertension and a greater proportion of people on higher incomes, compared to participants in the intervention arm. However, none of these observed differences were deemed to be sufficiently large to warrant adjustment in the relevant secondary analyses.

Sensitivity analyses checking for the impact of imbalances in randomisation, on the estimates for the primary outcome (difference in PHQ-9 scores at 3 months) using complete data only

| Model adjusted for imbalance | Estimate, 95% CI | *P* value |
| --- | --- | --- |
| 1. adjusted for gender, age-group, type of primary care centre, and baseline PHQ-9 scores | -0.64, (-1.75, 0.46) | 0.251 |
| 2. Model 1 + education group, hypertension, diabetes, baseline medication to treat depression, baseline EQ5D scores, baseline GAD-7 score | -0.30 (-1.51, 0.92) | 0.632 |

PHQ-9 scores range from 0 to 27, with higher scores representing more severe symptoms of depression

^a^ Difference in means between treatment arms were estimated using linear regression models, adjusted for stratified variables (gender, age-group, type of primary care centre (Family Health Strategy, traditional primary care centre, and baseline PHQ-9 score)

eTable 8: Comparison of baseline demographics and baseline measures for secondary outcomes, between trial arms, for participants followed-up at the three-month follow-up visit

| **Number/total sample size (%)** | | |
| --- | --- | --- |
|  | **Viva Vida (n=)** | **Enhanced usual care (n=)** |
| **Female, No. (%)** |  |  |
| **Age group, No. (%)** |  |  |
| 60-69 years |  |  |
| 70+ years |  |  |
| **Education, No. (%)** |  |  |
| None |  |  |
| 1-4 years |  |  |
| 5-8 years |  |  |
| >8 years |  |  |
| **Personal income, No. (%)** |  |  |
| Up to 1 MW |  |  |
| >1-2 MW |  |  |
| >2 MW |  |  |
| **Hypertension (self-reported), No (%)** |  |  |
| **Diabetes (self-reported), No (%)** |  |  |
| **Receiving pharmacological treatment for depression (self-reported), No (%)** |  |  |
| **Mean PHQ-9 score (SD)** |  |  |

*Abbreviations: MW: minimum wage (in 2021, the minimum wage in Brazil was BRL1110 (approximately US$213); PHQ-9: 9-item Patient Health Questionnaire for depression.*

eTable 9: Comparison of baseline demographics and secondary outcome measures, between trial arms, for participants followed-up at the five-month follow-up visit

| **Number/total sample size (%)** | | |
| --- | --- | --- |
| **Baseline variables** | **Viva Vida (n=)** | **Enhanced usual care (n=)** |
| **Female, No. (%)** |  |  |
| **Age group, No. (%)** |  |  |
| 60-69 years |  |  |
| 70+ years |  |  |
| **Education, No. (%)** |  |  |
| None |  |  |
| 1-4 years |  |  |
| 5-8 years |  |  |
| >8 years |  |  |
| **Personal income, No. (%)** |  |  |
| Up to 1 MW |  |  |
| >1-2 MW |  |  |
| >2 MW |  |  |
| **Hypertension (self-reported), No (%)** |  |  |
| **Diabetes (self-reported), No (%)** |  |  |
| **Receiving pharmacological treatment for depression (self-reported), No (%)** |  |  |
| **Mean PHQ-9 score (SD)** |  |  |

*Abbreviations: MW: minimum wage (in 2021, the minimum wage in Brazil was BRL1110 (approximately US$213); PHQ-9: 9-item Patient Health Questionnaire for depression.*

Moderation by pre-specified variables for either primary or secondary outcomes

None of the pre-specified subgroup analyses performed to investigate the potential differential intervention effects on the primary outcome of recovery was found to be relevant based on the significance of the corresponding likelihood ratio tests. eTable 13 shows the results of the likelihood ratio statistic for models with and without the interaction term at three months, and eTable 14 shows the results at five months. Multiplicative interactions were assessed on the log odds scale.

eTable 11: Results of Wald test for models testing for interaction terms with treatment allocations at three and five months with the outcome of mean difference in PHQ-9 scores

|  | ***P* Value of Wald test** | |
| --- | --- | --- |
| **Interaction term with treatment allocation** | **Three-month follow-up** | **Five-month follow-up** |
| Gender | 0.028 | 0.098 |
| Age | 0.429 | 0.657 |
| Education | 0.419 | 0.750 |
| Baseline PHQ-9 scores | 0.615 | 0.291 |
| Hypertension | 0.064 | 0.162 |
| Diabetes | 0.210 | 0.929 |
| Comorbid hypertension and diabetes | 0.167 | 0.336 |

*Abbreviations:* PHQ-9: 9-item Patient Health Questionnaire for depression.

**Table e12**

After running the regression model (fully adjusted) with interactions between gender and treatment allocation we need to interpret the coefficients.

| Variable | Coefficient | P value |
| --- | --- | --- |
| Gender | 2.41 (0.83, 4.00) | 0.003 |
| Treatment allocation | 0.86 (-0.96, 2.68) | 0.353 |
| Gender#Treatment allocation | -2.32 (-4.60, -0.05) | 0.045 |

1. The variable gender shows the difference in PHQ-9 scores between women and men in the control arm.
2. The variable arm shows the difference in PHQ 9 scores between the intervention and control arm, amongst men. This means there was a non-significant increase in PHQ-9 scores in men who received the intervention compared to mean who did not receive the intervention.
3. The difference in PHQ-9 scores between the intervention and control arms for women is -1.46 (-2.85, -0.08). This is the sum of the coefficients for treatment allocation 0.86 and the coefficient for the interaction term (-2.32). The 95% Cis are calculated using the lincom function in Stata. This means that women in the intervention arm have lower PHQ-9 scores than women in the control arm.

Sensitivity analyses checking for potential effect between mean number of elapsed days between baseline and the first follow-up visit

An additional analysis was performed to explore the potential effect of differences in the mean number of days elapsed from baseline to the first follow-up between the intervention arm (median number of days (MDN88)=, intra quartile range (IQR)87, 90) and control arm (MDN=88, IQR, 87, 90). Estimates from models using complete cases only, that adjusted for mean number of days, were very similar (Difference in mean PHQ-9 scores between the intervention and control arms at three months between intervention and control arm:-0.55; 95% CI:-1.66, 0.56 ), to the model that did not adjust for mean number of days (-0.62; -1.72, 0.49).
